# Reliable Online Auditory Cognitive Testing: An observational study

**DOI:** 10.1101/2024.09.17.24313794

**Authors:** Meher Lad, John-Paul Taylor, Tim D Griffiths

## Abstract

Technological advances have allowed researchers to conduct research remotely. Online auditory testing has received interest since the Covid-19 pandemic. A number of web-based developments have improved the range of auditory tasks during remote participation. Most of these studies have been conducted in young, motivated individuals who are comfortable with technology. Such studies have also used stimuli testing auditory perceptual abilities. Research on auditory cognitive abilities in real-world older adults is lacking.

In this study, we assess the reproducibility of a range of auditory cognitive abilities in older adults, with a range of hearing abilities, who took part in in-person and online experiments. Participants performed a questionnaire-based assessment and were asked to complete two verbal speech-in-noise perception tasks, for digits and sentences, and two auditory memory tasks, for different sound features. In the first part of the study, 58 Participants performed these tests in-person and online in order to test the reproducibility of the tasks. In the second part, 147 participants conducted all the tasks online in order to test if previously published findings from in-person research were reproducible.

We found that older adults under the age of 70 and those with a better hearing were more likely to take part in online testing. The questionnaire-based test had significantly better reproducibility than the behavioural auditory tests but there were no differences in reproducibility between in-person and online auditory cognitive metrics. Relationships between relationships with age and hearing thresholds in an in-person or online setting were not significantly different.

Furthermore, important relationships between auditory metrics, evidenced in literature previously, were reproducible online.

This study suggests that auditory cognitive testing may be reliably conducted online.

## Introduction

Online experimentation has allowed researchers to reach a large number of participants in a short time. This has increased the statistical power of studies and the reliability of their findings. Online recruitment platforms like Amazon Mechanical Turk and Prolific.co have become common venues for data collection, and stimulus creation and presentation is simplified through mediums like Gorilla, PsychoPy and Qualtrics [1,2]. This has resulted in a shift in research culture where an increasing number of laboratories are trying to answer their research questions by using these resources. The Covid-19 pandemic accelerated this process [3].

Online auditory testing has also increased during this time and a range of studies have evidenced the reliability and replicability of this method [4,5]. Auditory experiments rely on good controlled delivery of sounds with headphone usage, as the variability of background noise during sound presentation, equipment and choice of device used for presenting sounds can dramatically impact the user experience. For example, in an online setting it is not easy to verify whether a participant is using headphones, which attenuate background noise and ensure accurate delivery of sound requiring two audio channels, without visual verification. This introduces concerns regarding privacy. Many tests have been devised to overcome this problem [6]. This has led to a number of successful replications of auditory perceptual experiments [5,7]. However, a number of these studies have been conducted in young, motivated individuals who have volunteered for such experiments. People with severe hearing loss who may depend on hearing aids may not be able to physically accommodate headphones easily as listening without hearing aids may need to compensate for individual hearing impairments [8].

Some researchers have expressed concerns about the population being sampled from online databases [9]. Participants tend to be less representative of local populations. In-person research suffers from a lack of diversity in participation where people with limited literacy skills are often excluded from health research and this may be exacerbated when digital literacy is taken into consideration [10]. Older adults may be further marginalised although, it has been shown that web-based cognitive testing has been successfully implemented with older adults above the age of 65 [11]. Nevertheless, it is unclear what the effectiveness of online testing is on older adults with hearing difficulty.

There is a paucity of auditory cognitive research, using stimuli assessing speech-in-noise hearing for example, in older populations with a range of hearing abilities. Previous work has used verbal stimuli which are easier to perceive than simple or complex auditory stimuli below the level of speech [12]. We have learnt that participants are able to complete demanding tasks online with the particular experimental setup [13]. Learning effects of complex sound stimuli has also been studied in young and older adults recruited from Prolific.co successfully [14]. A combination of such stimuli with verbal stimuli have been studied in a similar participant group [15]. Whether reliable auditory cognitive online testing can be performed, using similar stimuli, in a ‘real-world’ population has not been previously attempted to our knowledge.

This first part of this exploratory study aimed to assess if there were population characteristics, such as age and hearing status, that were more likely to lead to online experimentation. We then assessed the reproducibility of auditory cognitive metrics in the participants who participated in-person and online. We asked participants to complete two verbal speech-in-noise perception tasks, for digits and sentences, and two auditory memory tasks, for different sound features. We compared whether participants scored similarly on a self-reported questionnaire about musical sophistication, the Goldsmiths Musical Sophistication Index (GMSI), compared with those obtained using computerised behavioural experiments for auditory cognition such as speech-in-noise measures and auditory memory [16]. We hypothesised better reproducibility of the questionnaire compared to the auditory cognitive tasks. We also assessed if there were differences in correlation coefficients between auditory measures and those that were shared across the two task settings, such as age and hearing thresholds.

For the second part of the study, we tested relationships between the auditory cognitive metrics that were identified previously by the in-person experimentation and were reproducible in an online group of participants. This included the participants from the first part and adults from the PREVENT study, a cohort of participants who were recruited to study risk-factors for dementia [17]. We wished to examine whether online auditory cognitive experiments would reproduce findings that we had found between GMSI and auditory memory for frequency precision, and speech-in-noise thresholds for sentence-in-babble perception and auditory memory [18,19].

During these in-person experiments stricter auditory experimental procedures were used and we expected participants to be more consistent, in this context, across in-person and online experiments compared to the computerised tests online.

## Methods

### Participants

153 older adults were recruited from a local Newcastle University volunteer database, the Join Dementia Research registry and from friends and family of people attending memory clinics for a dementia workup. They did not have a neurological or psychiatric condition at the time of participation. The age range of participants was 50 - 86 years with a mean of 67 years and a standard deviation of 10 years. 23 participants were active hearing aid users. All participants agreed to take part in an online follow-up assessment between 3 to 6 months after their initial in-person assessment. Participants who conducted the online assessments did not report any subjective change in their hearing abilities or any new medical conditions affecting their cognition. 58 of these participants took part in online repeat assessment. The performance of these participants was used to determine the reproducibility of auditory cognitive tests in-person vs. in an online setting. 89 additional participants were invited from another participant cohort, the PREVENT study. This study was originally developed to establish midlife risk factors for dementia in a multicentre UK cohort of participants. The aim of the online study was to test whether previously published findings could be replicated online. These were studies showing an association between auditory memory and sentence-in-noise perception ability, and auditory memory for frequency precision and musical sophistication [17–19]. The Newcastle and the PREVENT cohort were combined to provide a dataset to analyse online auditory cognitive metrics. Participants were compensated with £10 shopping vouchers for in-person testing and £5 vouchers for online experimentation.

The mean age of participants was 64.5 (±7.5) years of age. 57% of the participants were women. 5% of participants left school at the age of 15, 70% completed school and/or had a university degree and 24% had multiple degrees. 11% had normal hearing, 19% had mild hearing loss, 54% had moderate hearing loss and 16% had severe hearing loss. As shown in Figure 2, there were no participants above the age of 75 who participated online. Only 4 of 37 hearing aid users took part in the online experimentation session. Due to the small numbers of participants present in this group, further sub-group analysis was not conducted. In the study of online participants, across the Newcastle and PREVENT cohorts, the average age was 59 years (±7.7) years of age. The sample included 75 female and 47 male participants. As these participants did not have pure-tone audiometry performed, we were unable to grade the severity of hearing loss like for the in-person participants.

### Stimuli and Equipment

In-person testing was conducted in a soundproof room using a Dell Desktop computer with Sennheiser HD 201 circumaural headphones. An external sound card was used to process and deliver auditory stimuli and acoustic stimuli were presented at 70 dB A. All auditory cognitive tasks and the GMSI questionnaire, both of which are described in detail in the next section, were coded in Javascript and displayed with a Google Chrome web browser. The webpage was designed as a single-page application where all stimulus generation and user interaction was determined by processes that occurred ‘client-side’. The webpage hosted by Google Cloud Platform using Firebase Hosting. A Firebase Realtime Database was used to store anonymised results that were linked to each participant ID, which they used to access the testing portal at home at the follow-up assessment. At home, participants were instructed to use a desktop, laptop or tablet device with headphones and perform the test in a quiet room with no distractions. Hearing aid users were instructed to use over-ear headphones that do not interfere with the aids, if available, or to increase the computer device’s sound to a comfortable level before beginning the auditory tests.

### Experimental Procedure

Each participant had a 1-hour visit to the Auditory Laboratory at the Newcastle University Medical School. Pure tone audiometry (PTA) testing was performed on both ears from 250 Hz to 8 kHz at octave intervals for air conduction using an Interacoustics AS608e screening audiometer. Tones were manually presented as short bursts twice starting at 30 dB HL then increased in 5dB increments until comfortably audible if necessary. Then 5 dB HL reductions were made until the tone was not audible. This process was repeated twice and the lowest audible volume was chosen as the value for a particular frequency. If maximum amplification at 100 dB HLcould not be perceived then this was used as the ceiling value at a particular frequency. The overall mean of high frequency values between 4 to 8 kHz for the best ear was taken as the threshold value for an individual for further analysis. This value was chosen as high-frequency thresholds are suspected to deteriorate first in age-related hearing loss and previous research from our group has suggested that PTA thresholds in this range correlates with speech-in-noise difficulties [20,21]. Hearing status was determined as follows: normal if the mean threshold was below 20 dB HL, mild if between 20-40 dB HL, moderate if 40-60 dB HL hearing loss and severe if >80 dB HL.

The Digits-in-Noise (DIN) task involved participants listening to three digits on a background of speech-shaped white noise, created using white noise using the long-term average speech spectrum of speech stimuli between 80 to 10000 Hz. Participants had two practice trials at the beginning of the task to familiarise themselves with the stimuli at an SNR of 10dB. An adaptive 1-up, 1-down psychophysical paradigm was implemented whereby a correct response resulted in the SNR being reduced and an incorrect one caused the SNR to increase. The starting SNR was 0 dB and the step sizes decreased from 5 to 2 dB after 3 reversals, which then reduced to 0.5 dB after 3 more reversals. The run terminated after 10 reversals and the SNR at the last 5 reversals was averaged to calculate the DIN threshold for each participant. Lower SNR values indicated a better performance. As with the DIN task, participants had two practice trials at the beginning of the task to familiarise themselves with the stimuli at an SNR of 10 dB. The Speech-in-Babble (SIB) task consisted of participants listening to sentences on a background of 16-talker babble as described previously by our research group [18,21]. Target sentences had the form <NAME> <VERB> <NUMBER> <ADJECTIVE> <NOUN> (e.g. “Alan gives four pretty flowers”) and participants had to click on the correct word from a list of five columns (10 options for each word) shown on the screen with the same structure. The SIB threshold was determined using the same adaptive threshold procedure used in the DIN test. The starting parameters and adaptive design were exactly the same as the DIN task.

Auditory Memory (AuM) was tested using non-speech stimuli as previously described (Figure 1) (Lad et al., 2021). A one-second tone or AM modulated white noise stimulus was presented to a participant after which they were asked to ‘find’ the sound on a horizontal scale on a computer screen. Participants had to move a mouse and click on the line to produce a sound at that location. They could make as many clicks as they wanted with no set time limit. After they were satisfied with their choice they would advance to the next trial by pressing the ‘Enter’ key on a keyboard. Frequencies that determined the pure-tone sounds were chosen from a uniform distribution between 440-880 Hz and AM rates for the white noise stimulus were 5-20 Hz with a sinusoidal function used to apply this modulation. This parameter space, with an addition of 10% at either end of the scale, for each sound feature was mapped to the pixels of the horizontal scale on the screen during the matching phase. Hanning windows were applied to all synthetic sounds to avoid clicks and the beginning and end or the stimuli. The task consisted of 32 trials with the frequency and AM rate matching trials being interleaved. Participants had a short break after 16 trials. A Gaussian function was used to estimate the standard deviation of the errors in each trial across the whole experiment and the inverse of this value, the precision, was used for further analysis. Thus, one obtains a precision for frequency AuM (AuM (F)) and AM rate AuM (AuM (A)). Studies in vision have found that this measure better reflects the memory resource a participant can allocate in a given task [22]. Participants had two practice trials with each stimulus (2 for frequency and 2 for AM rate AuM) at the beginning of the task to familiarise themselves with the stimuli.

**Figure 1.**
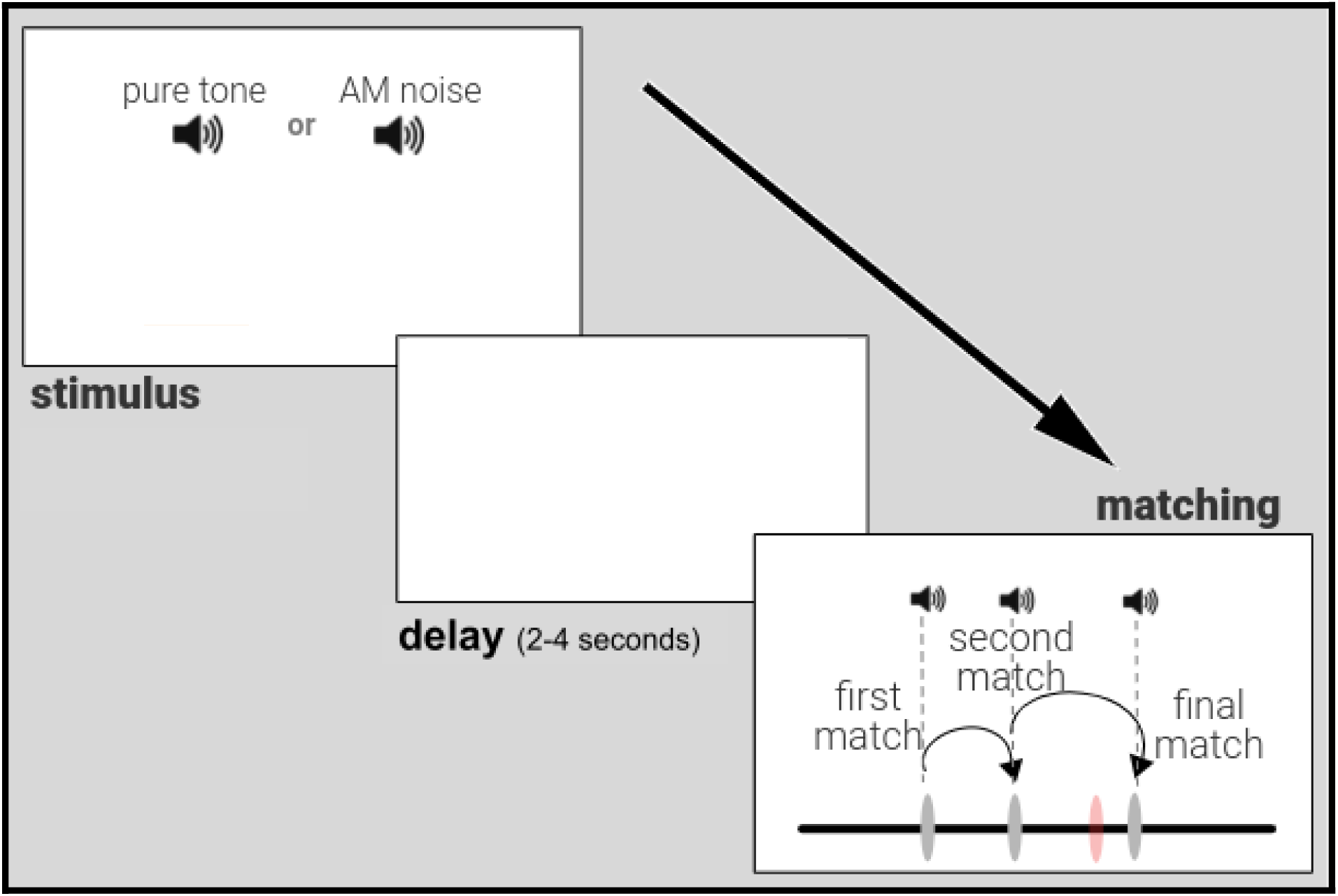
Auditory Memory Experiment. An auditory (pure tone or amplitude modulated noise) stimulus is presented for 1 second, then, after a delay of 2 to 4 seconds, participants can match sounds using a horizontal scale on the screen. The scale is linked to the parameter of interest (frequency for pure tone or AM rate) that can generate the original stimulus after exploring the parameter space to ‘find’ the stimulus. The figure shows an auditory matching trial where the participant’s ‘final match’ (rightmost dark grey marker on the scale) is shown in comparison to where the original stimulus (orange marker on the scale) is actually located. In this example, the participant first clicked on the scale to make a ‘first match’ (which produced a sound linked to the parameter at that location), then a ‘second match’ and then a ‘final match’. The discrepancy between the ‘final match’ location parameter and that of the original stimulus gives an ‘error’ for each trial that can be used to calculate the auditory working memory ‘precision’, the inverse of the standard deviation of errors from a trial target, for all auditory trials.

Finally, participants completed the short-version of the GMSI questionnaire consisting of 38 questions on paper as a test of musicality [16]. The GMSI is a self-report inventory that assesses individual differences in musical sophistication. It is said to measure the ability to engage with music in a comprehensive manner that does not just include musical instrumentation or training. For the purposes of this study, the GMSI was used as it has been previously used alongside the auditory cognitive measures in this study in an in-person setting [19]. We have previously shown a significant relationship between GMSI scores and auditory memory for frequency precision. This includes the total score of 266; the sum of the scores of the five domains of ‘Active Engagement’, ‘Perceptual Abilities’, ‘Musical Training’, ‘Singing Abilities’ and ‘Emotions’. This questionnaire was used as a control of internal consistency between in-person and online experimentation as we expected the change in testing modality to least affect a participant’s ability to answer multiple choice questions about their musical behaviours compared to the auditory tests.

### Statistical Analysis

AuM scores were log-transformed to achieve normal distributions. All other variables were normally distributed and converted to z-scores for further analysis.

Chi-square tests were used to calculate the difference in proportions in age groups and hearing groups between in-person and online participants. Pearson correlation coefficients were then used to measure the strength and direction of the linear relationship between auditory metrics. To evaluate the reproducibility of online versus in-person testing across different age groups, we used correlation coefficients and intraclass correlation coefficients. Steiger’s test was used to assess whether the correlation coefficients between in-person and online auditory metrics with shared variables such as age and PTA thresholds were significantly different.

The relationship between two variables while controlling for the effect of another variable (e.g. age) was performed after using linear regression to account for the influence of the control variable. This was used to assess the relationship between auditory cognitive metrics in-person then online. Finally, differences in correlation coefficients between these two sets of variables was calculated using bootstrapping. This was performed using repeated resampling of the data with replacement 1000 times. This allowed for the calculation of the mean difference and 95% confidence intervals for the correlation differences. Significant differences between correlation coefficients were established if the confidence intervals did not overlap. Due to the exploratory nature of this study, statistical correction for multiple corrections was not applied. All analyses were carried out using the Python programming language using Jupyter notebooks. SciPy and Pingouin packages were used.

## Results

### In-person vs. online auditory cognitive testing

Figure 2 shows the normalised age distribution of the in-person participant group and those that returned for online testing. The proportion of participants with different degrees of hearing loss is also shown. There was a significant difference in the proportion of participants over 70 years between the online and in-person groups, (χ2 = 9.40, p = 0.002), with a higher proportion of older participants in the in-person group. For the online participants, the proportion of those with moderate hearing loss was 54.05%, while the proportion with severe hearing loss was 16.22%. The mean hearing loss level was 45.84 ± 18.84 dB HL for online participants and 52.32 ± 23.03 dB HL for in-person participants. There was a statistically significant difference between the proportions of moderate and severe hearing loss categories among online participants (χ2 = 6.02,p = 0.014). For the in-person participants, the proportion of those with moderate hearing loss was 33.49%, and the proportion with severe hearing loss was 35.38%. There was no statistically significant difference in the proportions of moderate and severe hearing loss categories among in-person participants (χ2 = 1.07,p = 0.301).

**Figure 2.**
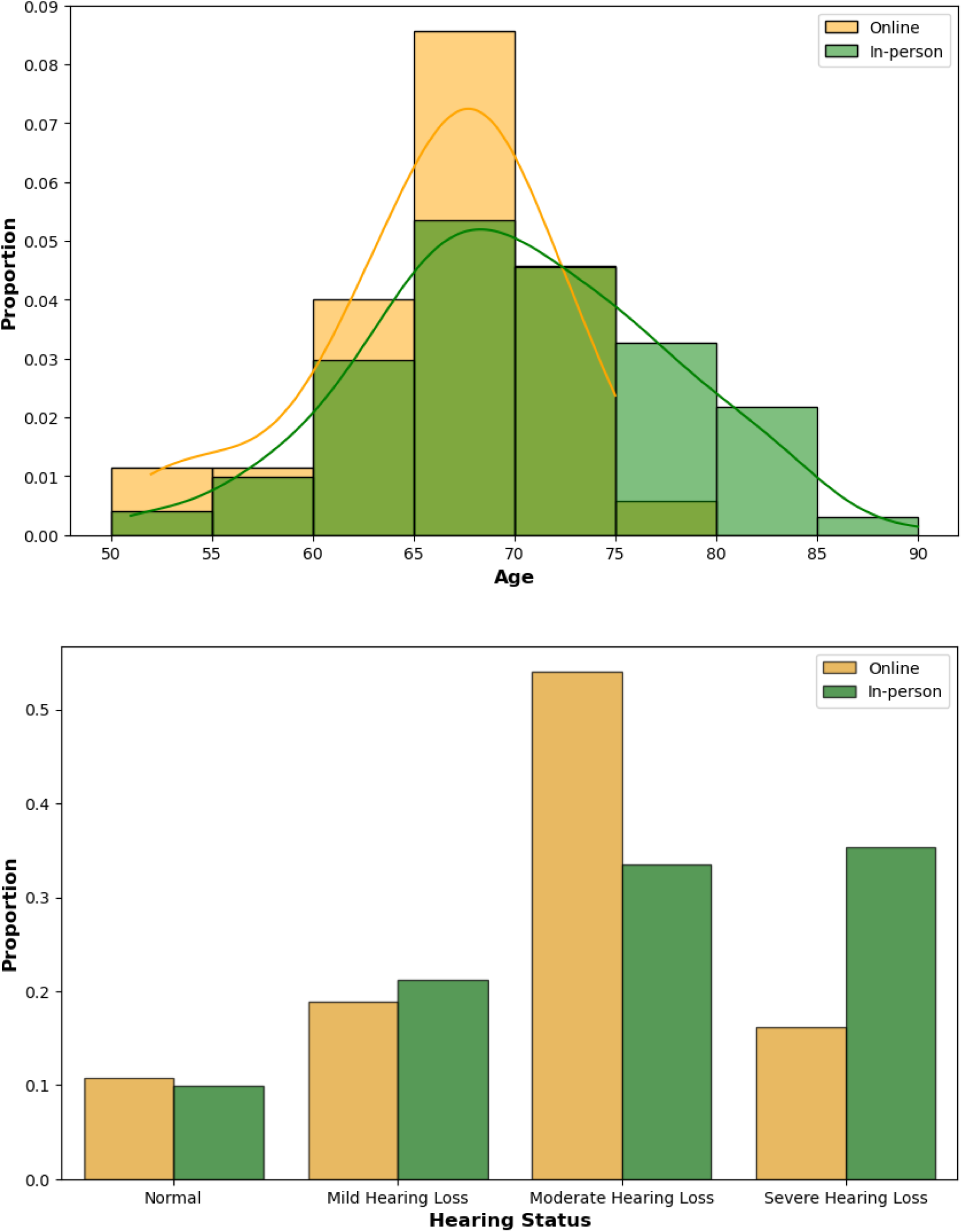
Age (top) and hearing status (bottom) in the in-person (N = 153) (green) vs online (N = 58) (range) cohort of participants. People above the age of 70 and those with severe hearing loss were less likely to participate in online auditory cognitive experimentation.

Table 1 shows the mean and standard deviation values of the auditory cognitive tests in both settings. There were no statistical differences between the mean values between in-person and online auditory measures. We assessed the reproducibility of various auditory and cognitive measures conducted in both lab and online settings using the Pearson’s correlation coefficient and intraclass correlation coefficients. The results indicated moderate reproducibility for DIN (r = 0.55, 95% CI [0.27, 0.74], p = 0.0005) and SIB (r = 0.55, 95% CI [0.27, 0.74], p = 0.0005). AuM (F) demonstrated good reliability (r = 0.75, 95% CI [0.56, 0.87], p < 0.0001), while AuM (A) showed lower reliability (r = 0.44, 95% CI [0.14, 0.67], p = 0.007). Notably, the GMSI exhibited the highest test-retest reliability among the measures (r = 0.82, 95% CI [0.67, 0.90], p < 0.0001).

**Table 1.**
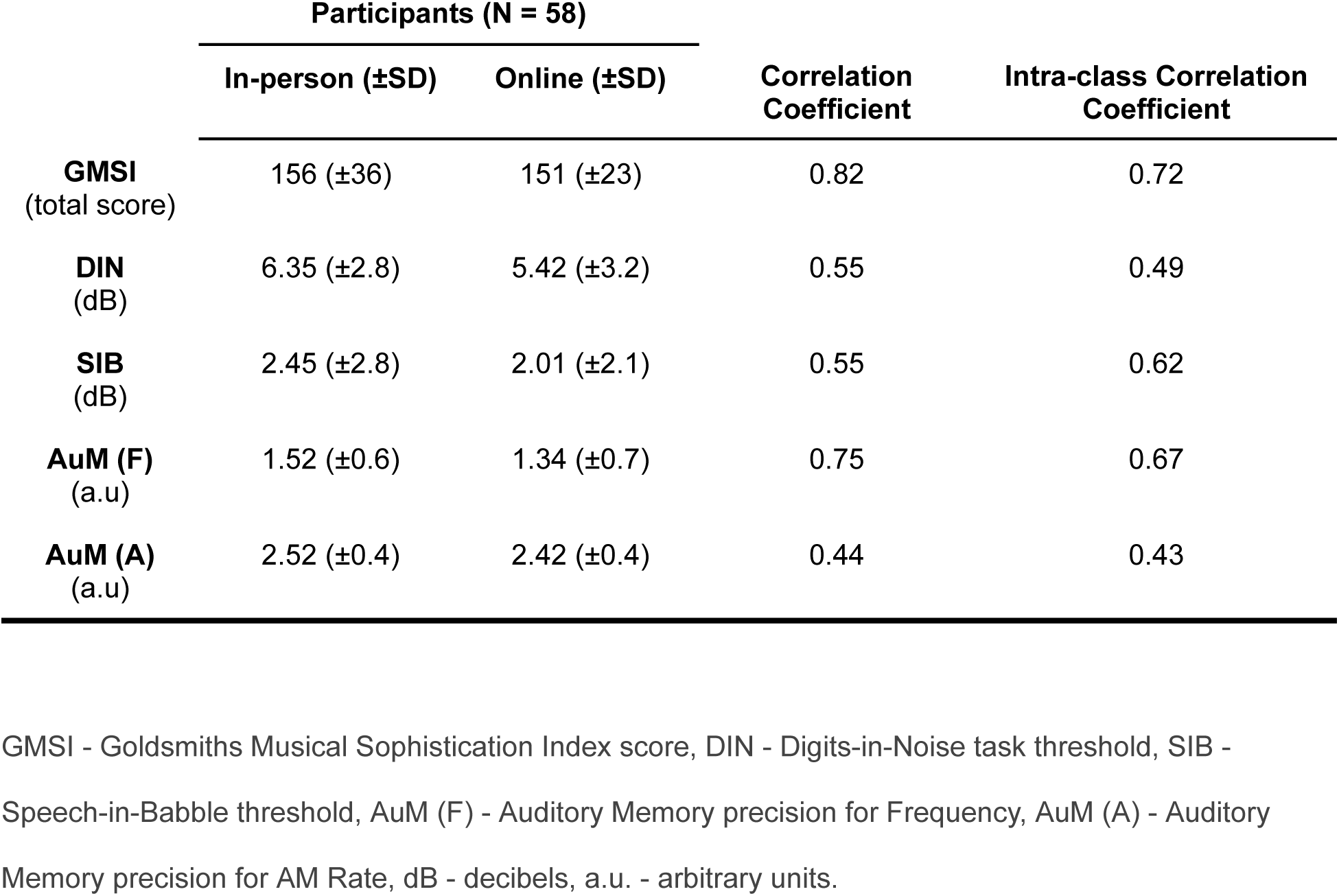
Auditory cognitive performance in-person and online.

|  | **Participants (N = 58)** |  | **Correlation Coefficient** | **Intra-class Correlation Coefficient** |
| --- | --- | --- | --- | --- |
|  | **In-person (±SD)** | **Online (±SD)** |  |  |
| **GMSI**<br>(total score) | 156 (±36) | 151 (±23) | 0.82 | 0.72 |
| **DIN**<br>(dB) | 6.35 (±2.8) | 5.42 (±3.2) | 0.55 | 0.49 |
| **SIB**<br>(dB) | 2.45 (±2.8) | 2.01 (±2.1) | 0.55 | 0.62 |
| **AuM (F)**<br>(a.u) | 1.52 (±0.6) | 1.34 (±0.7) | 0.75 | 0.67 |
| **AuM (A)**<br>(a.u) | 2.52 (±0.4) | 2.42 (±0.4) | 0.44 | 0.43 |

The reproducibility of the GMSI, with a correlation coefficient (R) of 0.82, was significantly greater than that of the SIB (mean difference = 0.25 (95% CI: 0.05, 0.46), p < 0.05) and AWM (A) (mean difference = 0.37 (95% CI: 0.08, 0.69), p < 0.05). The correlation coefficient of AWM (F) was not significantly higher than AWM (A) (mean difference = 0.31 (95% CI: -0.02, 0.67), p > 0.05) and the DIN was not significantly more reproducible than SIB (mean difference = -0.25 (95% CI: -0.36, 0.29), p > 0.05). The visualisations for these relationships are shown in Figure 3.

**Figure 3.**
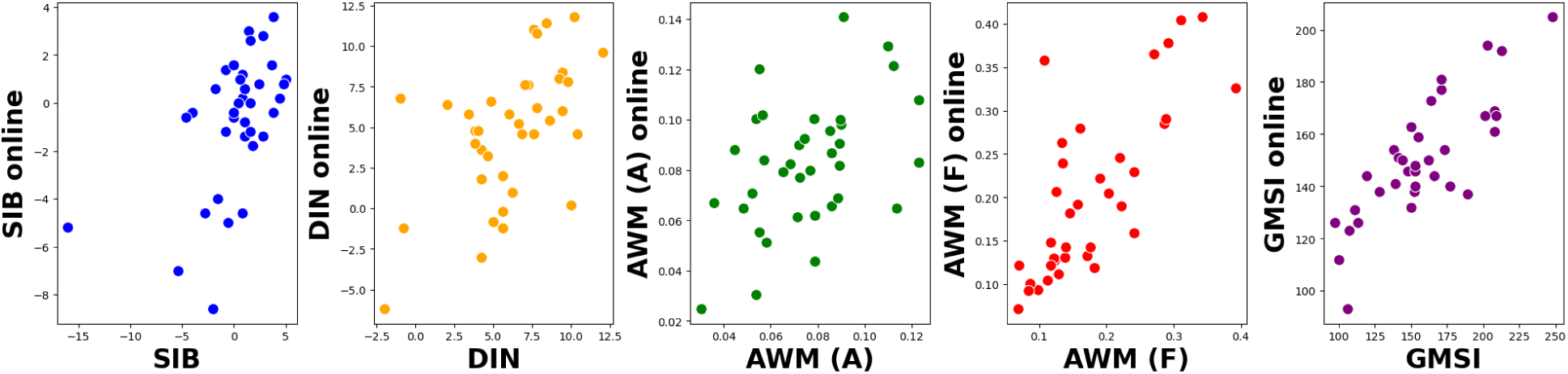
Participants’ (N = 58) data for various auditory cognitive measures and the GMSI questionnaire in an in-person and online setting. In-person data is shown on the x-axis whereas online metrics on the y-axis. GMSI - Goldsmiths Musical Sophistication Index score, DIN - Digits-in-Noise task threshold, SIB - Speech-in-Babble threshold, AuM (F) - Auditory Memory precision for Frequency, AuM (A) - Auditory Memory precision for AM Rate.

We tested whether there were differences in correlation coefficients between age and PTA thresholds, measured in person and auditory cognitive metrics measured in both settings using Steiger’s test. There were no significant differences between the correlations between age and any auditory metric in the online and in-person setting. Specifically, these include for the DIN (t = 0.165, p = 0.87), SIB (t = 0.162, p = 0.87), AuM (A) (t = -0.170, p = 0.87) and AuM (F) (t = 0.073, p = 0.94). Additionally, there were no significant differences between the correlations between PTA thresholds and any auditory metrics across settings. These include for the DIN (t = 0.005, p = 0.99), SIB (t = 0.031, p = 0.98), AuM (A) (t = -0.012, p = 0.99) and AuM (F) (t = 0.154, p = 0.88).

GMSI - Goldsmiths Musical Sophistication Index score, DIN - Digits-in-Noise task threshold, SIB - Speech-in-Babble threshold, AuM (F) - Auditory Memory precision for Frequency, AuM (A) - Auditory Memory precision for AM Rate, dB - decibels, a.u. - arbitrary units.

### Online auditory cognitive testing associations

We tested whether auditory cognitive associations between variables obtained in-person were reproducible online with a larger group of online participants. The latter included the addition of a new group of participants, from the PREVENT study, who had not previously performed the task. This was done to improve the statistical power of the online findings. We examined the reproducibility of the association between AuM for frequency and GMSI scores and between SIB and AuM, when conducted in-person and online. This has been studied previously by our group on an in-person participant group [18,19].

The correlation coefficients for the relationship between SIB and AuM (A) were statistically significant for in-person testing (R = 0.46, 95% CI [0.34, 0.56], p < 0.001) and online testing (R = 0.18, 95% CI [0.0, 0.34], p < 0.05) (Figure 4). Bootstrapping was used to generate 95% confidence intervals for the correlation coefficients to test if they were significantly different at the significance level with an alpha of 0.05. We did not find any differences across the two settings for this relationship. Similarly, the correlation coefficients for the relationship between GMSI scores and AuM (F) were statistically significant for in-person testing (R = 0.52, 95% CI [0.42, 0.61], p < 0.001) and online testing (R = 0.47, 95% CI [0.32, 0.6], p < 0.001). We did not find any differences across the two settings for this relationship. This suggested that there were no differences in the correlation of these auditory measures in an in-person or online setting.

**Figure 4.**
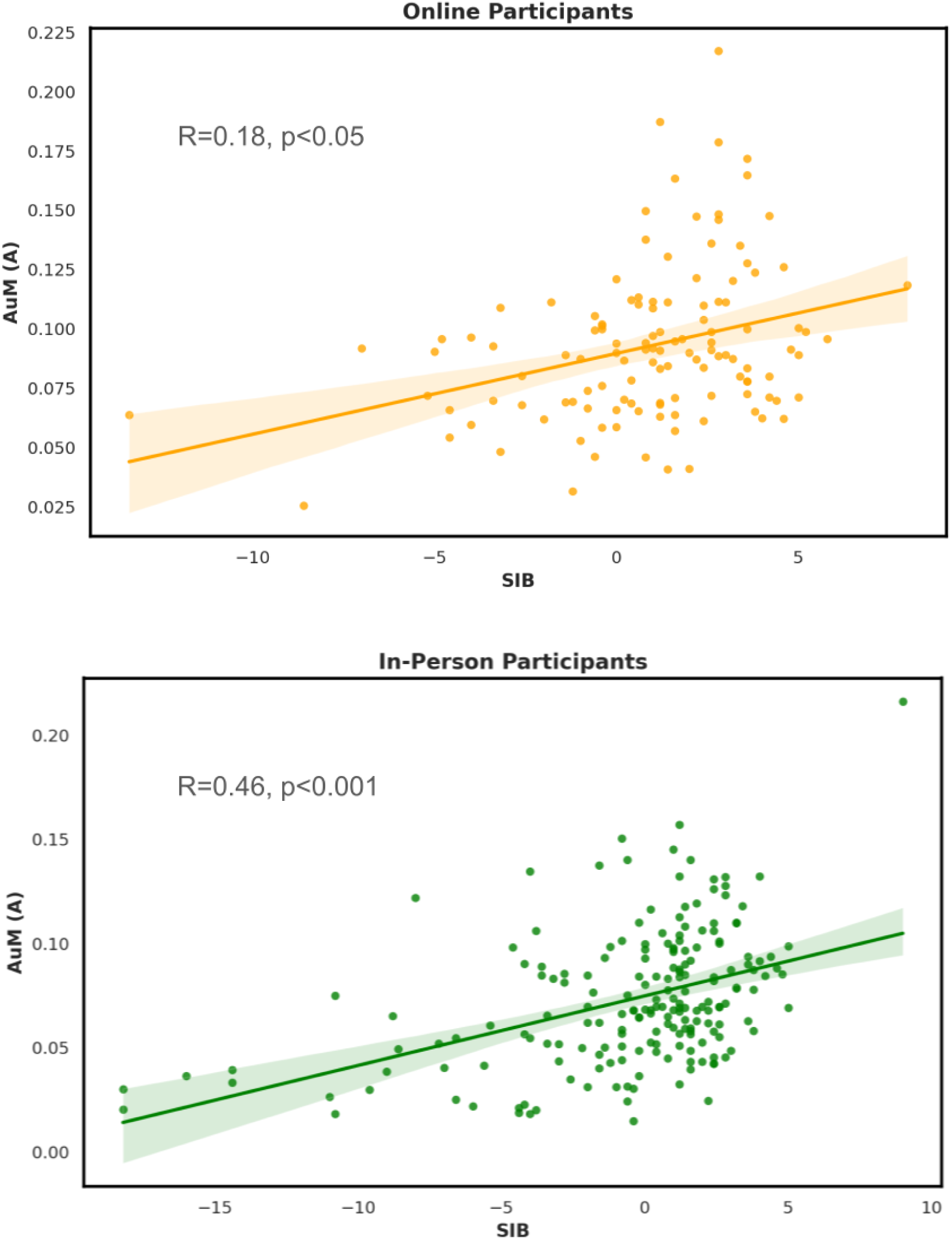
The relationship between sentence-in-babble (SIB) perception ability and auditory memory for amplitude modulation rate (AuM (A)) in online (N = 147) (top panel - orange) and in-person (N = 153) (bottom panel - green) participants. Shaded areas indicate confidence intervals for the regression lines of best fit. There is greater variability for online participant data compared to in-person data.

## Discussion

The principal findings of this study were that remote auditory testing of cognition is reliable when conducted online. However, online participation may not be fully representative of the participants who may take part in-person. We found that older participants and those with severe hearing loss were less likely to participate online. Furthermore, we found that the questionnaire-based test, the GMSI, had the best reproducibility as compared to the auditory cognitive tasks, which did not differ significantly when conducted in-person or online. Importantly, previously published findings were reproducible online suggesting that online testing may be a suitable avenue to test auditory cognition.

Although this study has not been conducted previously, our experience with online auditory experimentation was similar to previous work in the field [5,14,15]. Older adults in the ‘real-world’ were able to successfully negotiate a self-directed online auditory experimentation portal and give reliable results in comparison to in-person testing. Our study differed from the previous work in the type and range of stimuli across the same experimental paradigm. For example, we tested the perception of digits and sentences on a noisy background and sounds manipulated by frequency and temporal fluctuations. This allowed us to test different types of auditory stimuli across the same type of experiment (speech-in-noise perception and auditory memory). We did not find any differences in the reproducibility of any of these measures across in-person or online settings. Importantly, there was no significant difference when comparing the relationships of these metrics with shared variables across both settings such as age and hearing thresholds.

A focus of this study was to engage participants with a range of hearing abilities that are recruited for in-person studies. Despite an increase in online research methods, there may be an underrepresentation of participants who lack adequate digital literacy or find it difficult to interact with computer-generated stimuli. In particular, people with hearing loss may rely more on visual cues to understand information, which may be absent from typical auditory research performed on healthy young-adults without hearing difficulty [23]. Although we did not explore the qualitative reasons behind the reduced proportion of participants with severe hearing loss in our online cohort, these participants were also more likely to be hearing aid users, which could have interfered with their use of headphones. Data on whether participants were using bluetooth enabled devices which allowed connection to their electronic devices was not collected.

Additionally, to maximise participation in the study we gave volunteers up to 6 months to complete their online experiment. Although this may have improved participant retention to a degree, we did not re-measure their pure-tone audiometric thresholds to ensure the peripheral hearing was the same as in the in-person experiment. This may have improved the reproducibility of the online auditory cognitive metrics.

We also found no differences in reproducibility of auditory cognitive stimuli with particular sound parameters. There could have been better reproducibility for AuM for frequency due to the invariance of sounds with a pitch to changes in timbre over different electronic devices [24]. The pitch of a sound is also less likely to be affected if it is played on headphones or speakers and due to the sound cards in electronic devices. For example, the frequency space from which the pure-tones were generated were from 440 to 880 Hz which is captured by most audio devices. Speech-based sounds and the amplitude modulated white noise stimuli would have only been limited by the bandwidth of the electronic devices and equipment. This could have affected the performance in the speech-in-noise tasks and AuM (A) task online and contributed to the lower correlation coefficient. However, these were not statistically different. Further work is needed to clarify the reasons for this.

The observed similarities between amplitude-modulated noise stimuli and speech in conversations highlight their importance in understanding auditory processing, particularly in relation to speech-in-noise perception ability. Both modalities exhibit temporal amplitude fluctuations critical for investigating auditory system functions without background noise [25]. Modulated noise mimics speech’s natural rhythms, aiding in the study of temporal processing in speech. Cognitive engagement is essential in both, requiring attention and working memory to decode or track fluctuations, however, the presence of linguistic and phonetic information in speech is not found in the generated sounds of the AuM task. Despite these differences, the correlation between non-speech stimuli in auditory modulation tasks and verbal SIN tasks indicates underlying auditory cognitive abilities shared across both domains, bridging the gap between speech and non-speech auditory processing. This suggests a broader scope of auditory cognition beyond specific linguistic content, highlighting fundamental auditory processing mechanisms applicable across different auditory stimuli. The reproducibility of this relationship across the in-person and online settings suggests that these fundamental connections between the stimuli can be investigated online.

The home environment of a participant may affect a participant’s performance in a number of ways. Although there were instructions to set the volume level to one that was comfortable in our task, participants may alter this through the course of the tasks. The home environment is also prone to background fluctuations in noise that may not be observed in strict laboratory conditions. These interactions may affect the listening ability of participants despite the sounds being at a comfortable level [26]. Finally, despite using headphones, the sound quality that may have been transmitted through them could have varied between participants resulting in any changes in sound quality mentioned above. It was impossible to monitor this accurately during this study. Despite all of these challenges, it is promising for the auditory measures used in this study to display reproducibility across a range of different speech-in-noise stimuli and auditory memory tasks with different sound features. However, the participants in this study were recruited from a local university volunteer database, which may introduce a selection bias as these individuals are more likely to have prior experience with research participation. Therefore, while our findings provide valuable insights, they may not be fully generalisable to the broader population and this limitation should be considered when interpreting the results.

The strengths of this study include inclusion of ‘real-world’ older adult participants in independent cohorts, with a range of hearing abilities, to perform in-person and online auditory experiments. This allowed us to accurately assess the differences in within-subject parameters that are attributable to online testing. We also used multiple types of verbal and non-verbal stimuli to test auditory cognitive abilities which allowed us to find differences in performance for particular sound features, like frequency vs. amplitude modulation, in both settings.

Future work could include an assessment of the reasons for which participants did not participate in the online session despite agreeing to participate in this initially. There may have been other factors, rather than those related to hearing, that influenced these decisions for each participant. Other improvements to the study design include remote guided testing where the participant performs the task on their home computer but through a screen share [27]. This would ensure that the home environment and equipment could ‘pass’ a minimum standard to perform the tasks. Recruitment of a larger group of participants who return for online testing may allow further analysis to assess the reproducibility of the auditory measures across different age groups and hearing loss severity levels. This will allow us to assess if these are factors that affect the reproducibility of certain results.

## Data Availability

Data and analysis code is available from OSF.

## Acknowledgements

The Oxford C NHS Research Ethics Committee (21/SC/0139) approved this study on the 12th of June 2021. All participants gave written consent to participate and publish data. Data and analysis code is available from OSF. The Newcastle NIHR Biomedical Research Centre supported the study. ML has received funding from the National Institute of Health Research, Alzheimer’s Research UK, the Guarantors of Brain and Medical Research Council (MR/V006568/1) in the United Kingdom. JT receives funding from the National Institute of Health Research Biomedical Research Centre, United Kingdom in Newcastle upon Tyne. TG receives funding from the Medical Research Council (MR/T032553/1), Wellcome Trust (WT106964MA) and National Institute on Deafness and Other Communication Disorders (DC000242 36).

## Acknowledgements

The authors have no conflicts of interest to declare.

## References

1. Sauter M, Draschkow D, Mack W. Building, Hosting and Recruiting: A Brief Introduction to Running Behavioral Experiments Online. Brain Sci Multidisciplinary Digital Publishing Institute; 2020 Apr;10(4):251. doi: 10.3390/brainsci10040251

2. Tomczak J, Gordon A, Adams J, Pickering JS, Hodges N, Evershed JK. What over 1,000,000 participants tell us about online research protocols. Front Hum Neurosci 2023;17. Available from: https://www.frontiersin.org/articles/10.3389/fnhum.2023.1228365 [accessed Jan 26, 2024]

3. Peyton K, Huber GA, Coppock A. The Generalizability of Online Experiments Conducted During the COVID-19 Pandemic. J Exp Polit Sci 2022 Nov;9(3):379–394. doi: 10.1017/XPS.2021.17

4. Eerola T, Armitage J, Lavan N, Knight S. Online Data Collection in Auditory Perception and Cognition Research: Recruitment, Testing, Data Quality and Ethical Considerations. Audit Percept Cogn Routledge; 2021 Oct 2;4(3–4):251–280. doi: 10.1080/25742442.2021.2007718

5. Zhao S, Brown CA, Holt LL, Dick F. Robust and Efficient Online Auditory Psychophysics. Trends Hear SAGE Publications Inc; 2022 Jan 1;26:23312165221118792. doi: 10.1177/23312165221118792

6. Milne AE, Bianco R, Poole KC, Zhao S, Oxenham AJ, Billig AJ, Chait M. An online headphone screening test based on dichotic pitch. Behav Res Methods 2021 Aug 1;53(4):1551–1562. doi: 10.3758/s13428-020-01514-0

7. Mok BA, Viswanathan V, Borjigin A, Singh R, Kafi H, Bharadwaj HM. Web-based Psychoacoustics: Hearing Screening, Infrastructure, and Validation. bioRxiv; 2021.p. 2021.05.10.443520. doi: 10.1101/2021.05.10.443520

8. Baumgartner H, Schulz A, Hein A, Holube I, Herzke T. A fitting method for headphones to compensate individual hearing impairments. 2009 3rd Int Conf Pervasive Comput Technol Healthc 2009. p. 1–8. doi: 10.4108/ICST.PERVASIVEHEALTH2009.5991

9. Stewart N, Chandler J, Paolacci G. Crowdsourcing Samples in Cognitive Science. Trends Cogn Sci 2017 Oct 1;21(10):736–748. doi: 10.1016/j.tics.2017.06.007

10. Loignon C, Dupéré S, Leblanc C, Truchon K, Bouchard A, Arsenault J, Pinheiro Carvalho J, Boudreault-Fournier A, Marcotte SA. Equity and inclusivity in research: co-creation of a digital platform with representatives of marginalized populations to enhance the involvement in research of people with limited literacy skills. Res Involv Engagem 2021 Oct 5;7(1):70. doi: 10.1186/s40900-021-00313-x

11. Cyr A-A, Romero K, Galin-Corini L. Web-Based Cognitive Testing of Older Adults in Person Versus at Home: Within-Subjects Comparison Study. JMIR Aging 2021 Feb 1;4(1):e23384. doi: 10.2196/23384

12. Slote J, Strand JF. Conducting spoken word recognition research online: Validation and a new timing method. Behav Res Methods 2016 Jun 1;48(2):553–566. doi: 10.3758/s13428-015-0599-7

13. Bianco R, Mills G, de Kerangal M, Rosen S, Chait M. Reward Enhances Online Participants’ Engagement With a Demanding Auditory Task. Trends Hear SAGE Publications Inc; 2021 Jan 1;25:23312165211025941. doi: 10.1177/23312165211025941

14. Bianco R, Hall ETR, Pearce MT, Chait M. Implicit auditory memory in older listeners: From encoding to 6-month retention. Curr Res Neurobiol 2023 Jan 1;5:100115. doi: 10.1016/j.crneur.2023.100115

15. Bianco R, Chait M. No Link Between Speech-in-Noise Perception and Auditory Sensory Memory - Evidence From a Large Cohort of Older and Younger Listeners. Trends Hear 2023;27:23312165231190688. PMID:37828868

16. Müllensiefen D, Gingras B, Musil J, Stewart L. The Musicality of Non-Musicians: An Index for Assessing Musical Sophistication in the General Population. PLOS ONE Public Library of Science; 2014 Feb 26;9(2):e89642. doi: 10.1371/journal.pone.0089642

17. Ritchie CW, Ritchie K. The PREVENT study: a prospective cohort study to identify mid-life biomarkers of late-onset Alzheimer’s disease. BMJ Open British Medical Journal Publishing Group; 2012 Jan 1;2(6):e001893. PMID:23166135

18. Lad M, Holmes E, Chu A, Griffiths TD. Speech-in-noise detection is related to auditory working memory precision for frequency. Sci Rep 2020 19;10(1):13997. PMID:32814792

19. Lad M, Billig AJ, Kumar S, Griffiths TD. A specific relationship between musical sophistication and auditory working memory. 2021 Jul p. 2021.07.08.451659. doi: 10.1101/2021.07.08.451659

20. Tl W, R C, L C, Dm N, Kj C. Changes in hearing thresholds over 10 years in older adults. J Am Acad Audiol J Am Acad Audiol; 2008 Apr;19(4). PMID:18795468

21. Holmes E, Griffiths TD. ‘Normal’ hearing thresholds and fundamental auditory grouping processes predict difficulties with speech-in-noise perception. Sci Rep 2019 Nov 14;9(1):1–11. doi: 10.1038/s41598-019-53353-5

22. Ma WJ, Husain M, Bays PM. Changing concepts of working memory. Nat Neurosci 2014 Mar;17(3):347–356. PMID:24569831

23. Suess N, Hauswald A, Zehentner V, Depireux J, Herzog G, Rösch S, Weisz N. Influence of linguistic properties and hearing impairment on visual speech perception skills in the German language. PLoS ONE 2022 Sep 30;17(9):e0275585. PMID:36178907

24. McPherson MJ, McDermott JH. Relative pitch representations and invariance to timbre. Cognition 2023 Mar 1;232:105327. doi: 10.1016/j.cognition.2022.105327

25. Goswami U. Speech rhythm and language acquisition: an amplitude modulation phase hierarchy perspective. Ann N Y Acad Sci 2019;1453(1):67–78. doi: 10.1111/nyas.14137

26. Ruggles D, Bharadwaj H, Shinn-Cunningham BG. Normal hearing is not enough to guarantee robust encoding of suprathreshold features important in everyday communication. Proc Natl Acad Sci Proceedings of the National Academy of Sciences; 2011 Sep 13;108(37):15516–15521. doi: 10.1073/pnas.1108912108

27. Leong V, Raheel K, Sim JY, Kacker K, Karlaftis VM, Vassiliu C, Kalaivanan K, Chen SHA, Robbins TW, Sahakian BJ, Kourtzi Z. A New Remote Guided Method for Supervised Web-Based Cognitive Testing to Ensure High-Quality Data: Development and Usability Study. J Med Internet Res 2022 Jan 6;24(1):e28368. doi: 10.2196/28368

